# Immunogenic superiority and safety of Biological E’s CORBEVAX™ vaccine compared to COVISHIELD™ (ChAdOx1 nCoV-19) vaccine studied in a phase III, single blind, multicenter, randomized clinical trial

**DOI:** 10.1101/2022.03.20.22271891

**Authors:** Subhash Thuluva, Vikram Paradkar, Kishore Turaga, SubbaReddy Gunneri, Vijay Yerroju, Rammohan Mogulla, PV Suneetha, Mahesh Kyasani, Senthil Kumar Manoharan, Srikanth Adabala, Aditya Sri Javvadi, Guruprasad Medigeshi, Janmejay Singh, Heena Shaman, Akshay Binayke, Aymaan Zaheer, Amit Awasthi, Chandramani Singh, A Venkateshwar Rao, Indranil Basu, Khobragade Akash Ashok Kumar, Anil Kumar Pandey

## Abstract

**Background:** Optimum formulation of Biological E’s CORBEVAX™ vaccine that contains protein sub unit of Receptor Binding Domain (RBD) from the spike protein of SARS-COV-2 formulated with aluminum hydroxide (Al3+) and CpG1018 as adjuvants was selected in phase-1 and 2 studies and proven to be safe, well tolerated and immunogenic in healthy adult population. In the current study, additional data was generated to determine immunogenic superiority of CORBEVAX™ vaccine over COVISHIELD™ vaccine and safety in larger and older population.

**Methods:** This is a phase III prospective, single blinded, randomized, active controlled study (CTRI/2021/08/036074) conducted at 20 sites across India in healthy adults aged between 18-80 years. This study has two arms; immunogenicity arm and safety arm. Participants in immunogenicity arm were randomized equally to either CORBEVAX™™ or COVISHIELD™ vaccination groups to determine the immunogenic superiority. Healthy adults without a history of Covid-19 vaccination or SARS-CoV-2 infection, were enrolled.

**Findings:** The safety profile of CORBEVAX™ vaccine was comparable to the comparator vaccine COVISHIELD™ in terms of overall AE rates, related AE rates and medically attended AEs. Majority of reported AEs were mild in nature, and overall CORBEVAX™ appeared to cause fewer local and systemic adverse reactions/events. Overall, two grade-3 serious AEs (Dengue fever and femur fracture) were reported and they are unrelated to study vaccine. Neutralizing Antibody titers, against both Ancestral and Delta strain, induced post two-dose vaccination regimen were higher in the CORBEVAX™ arm as compared to COVISHIELD™ and the analysis of GMT ratios demonstrated immunogenic superiority of CORBEVAX™ in comparison with COVISHIELD™. Both CORBEVAX™ and COVISHIELD™ vaccines showed comparable seroconversion post vaccination when assessed against anti-RBD IgG response. The subjects in CORBEVAX™ cohort also exhibited higher Interferon-gamma secreting PBMC’s post stimulation with SARS-COV-2 RBD peptides than the subjects in COVISHIELD™ cohort.

**Interpretations:** Neutralizing antibody titers induced by CORBEVAX™ vaccine against Delta and Ancestral strains were protective, indicative of vaccine effectiveness of >90% for prevention of symptomatic infections based on the Correlates of Protection assessment performed during Moderna and Astra-Zeneca vaccine Phase III studies. Safety findings revealed that CORBEVAX™ vaccine has excellent safety profile when tested in larger and older population.

**Funding:** BIRAC-division of Department of Biotechnology, Government of India, and the Coalition for Epidemic Preparedness Innovations funded the study.

## INTRODUCTION

Severe acute respiratory syndrome coronavirus 2 (SARS-CoV-2) infections has led to a global COVID-19 pandemic (WHO Covid-19 Situation report – 51) and there has been widespread impact on health, including substantial mortality among older people and those with pre-existing health conditions.^1^ It also severely affected global economy. Vaccines play an important role in increasing population immunity and preventing severe form of the disease. Global efforts to develop and test vaccines against SARS-CoV2 has resulted in approval of several vaccine candidates with varied efficacy.^2^ WHO has so far granted emergency use listing to ten covid-19 vaccines and five others are under assessment. Out of ten WHO recognized vaccines, only Novavax COVID-19 Vaccine (NVX-CoV2373) is a subunit vaccine and all others are either inactivated virus, nucleic acid based or viral vector-based vaccines. NVX-CoV2373 is based on of full-length, pre-fusion trimers of spike glycoprotein of prototype Wuhan sequence. Biological E developed a protein sub-unit vaccine (known as CORBEVAX™) that consists of Receptor Binding Domain (RBD) from the spike protein of SARS-COV-2 as the antigen that is formulated with aluminum hydroxide (Al3+) and CpG1018 as adjuvants after preclinical evaluation.^3,4^ The recombinant protein is expressed in yeast and uses a technology similar to the one for producing the recombinant hepatitis B vaccine that has been widely accepted for decades by populations in low- and middle-income countries. Therefore, CORBEVAX™ is potentially well suited as a COVID-19 vaccine for global health and to address both vaccine equity and hesitancy.

The optimum dose of the candidate vaccine, CORBEVAX™, was determined in Phase I/II studies conducted in adults and consists of 25 mcg of RBD protein, 750 mcg of Al3+ (in Aluminum Hydroxide) and 750 mcg of CpG1018 per 0·5 mL dose. The optimum formulation showed excellent safety profile with minimal reactogenicity and high humoral immune response in terms of anti-RBD IgG titers and neutralizing antibody titers against Ancestral, Beta and Delta strains of SARS-COV-2 as well as desired Th1 skew of the cellular immune response.^5^ In the current phase III study using the optimum formulation of CORBEVAX™, we report the safety and immunogenic superiority of CORBEVAX™ vaccine over COVISHIELD™ vaccine.

## METHODS

### Study Design and Study Population

This is an ongoing phase III prospective, single blinded, randomized, active controlled study conducted at 18 sites across India in accordance with the principles defined in the Declaration of Helsinki, International Conference on Harmonization guidelines (Good Clinical Practices), and the local regulatory guidelines. The Investigational Review Board or Ethics Committee at each study site approved the protocol. All participants provided written informed consent before enrollment into the study. Participants were healthy adults, aged between 18 to 80 years. This study has two arms; one is to determine the immunogenic superiority of CORBEVAX™ vaccine over COVISHIELD™ vaccine. Other arm is to determine only safety of CORBEVAX™ vaccine. Subjects enrolled into immunogenicity arm were also assessed for safety. A total of 6485 subjects were screened, of which 2140 subjects randomized and 2139 subjects enrolled (639 subjects in immunogenicity arm & 1500 subjects in safety arm). Safety data until day 56 (safety) and immunogenicity data at day 42 compared to day 0 (baseline) is presented in this manuscript.

Participants were seronegative to anti-SARS-CoV-2 IgG antibody prior to randomization into immunogenicity arm, whereas in safety arm subjects were randomized irrespective of their serostatus for SARS-CoV-2. Other key eligibility criteria applicable to all participants were: virologically negative to SARS-CoV-2 infection confirmed by RT-PCR test, seronegative to HIV 1 & 2, HBV and HCV infection. Health status assessed during the screening period was based on medical history and clinical laboratory findings, vital signs, and physical examination. All those who were with axillary temperature of more than 38·0°C, part of any other clinical trial, with a hhistory of vaccination with any investigational vaccine against Covid-19 disease, known allergy to vaccine components, or were on immunosuppressants, immunodeficient conditions were excluded from the study. Complete list of eligibility criteria provided as supplementary information.

During the conduct of this study, there were no major protocol deviations reported at any of the study sites. Few subjects reported for their visits out of window period but these deviations were not found to be significant and all deviations were notified to ethics committees of the respective study sites.

### Randomization and masking

Participants enrolled into immunogenicity arm were randomized equally either to receive CORBEVAX™™ vaccine or COVISHIELD™ vaccine. Randomization occurred after all screening-related activities were completed and prior to the first dose of study vaccine using the interactive web response system (IWRS) platform. A subject was considered randomized when he/she has met all the eligibility criteria and have received the randomization number from IWRS. A randomization scheme was generated by using a validated system. This is also a single-blind study where study participants randomized into immunogenicity arm are kept blinded of the vaccination group to which they have been assigned, but the investigator and study staff are aware of the assigned group (CORBEVAX™ or COVISHIELD™).

### Procedure

Biological E’s CORBEVAX™ vaccine is based on recombinant RBD protein, which is produced in Pichia Pastoris culture as secretory protein and purified via multiple chromatography and ultrafiltration/normal-filtration steps. The RBD subunit is co-formulated along with aluminum Hydroxide and CpG1018.^5^

The active comparator used in immunogenicity arm of the study is COVISHIELD™ (ChAdOx1 nCoV-19) is a Covid-19 vaccine. This vaccine is based on recombinant, replication-deficient chimpanzee adenovirus vector encoding the SARS-CoV-2 Spike (S) glycoprotein, produced in genetically modified human embryonic kidney (HEK) 293 cells (CDSCO.gov). In India, it is manufactured by Serum Institute of India and approved to active immunization of individuals ≥18 years old for the prevention of COVID-19.

A 0·5 mL dose of the candidate COVID-19 vaccine (CORBEVAX™) or COVISHIELD™ vaccine was administered via an intramuscular (IM) injection into the deltoid muscle of the non-dominant arm in a 2-dose schedule with 28 days’ interval between doses. No prophylactic medication was prescribed either before or after vaccination. Follow-up were scheduled at day 42, day 56, day 118 (3 months post second dose) and day 208 (6 months post second dose). Study is ongoing and subjects are under follow-up period.

Participants were evaluated with SARS-CoV-2 real-time RT-PCR for absence of infection using Diasorin kit and a serology test for seronegative status (Anti-SARS CoV-2 Human S1/S2 IgG ELISA COVID using Diasorin kit).^6^ Participants who were negative for both Anti-SARS CoV-2 human S1/S2 IgG antibodies and SARS CoV-2 infection were enrolled to study immune responses (immunogenicity arm).

### Outcomes

The primary outcome of the study was demonstration of immunogenic superiority of BE’s CORBEVAX™ vaccine against COVISHIELD™ vaccine in terms of GMTs of anti-SARS-CoV-2 virus neutralizing antibodies at day 42 (14 days after 2nd dose). Other secondary outcomes were demonstration of immune response against the Delta variant in terms virus neutralizing antibodies (VNA) at day-42. Anti-RBD antibody concentration in terms of GMC’s and to descriptively assess the safety, tolerability and reactogenicity of CORBEVAX™ vaccine during the study period. The exploratory end-point included cellular immune response assessment in a subset of subjects via ELISPOT method.

### Safety assessments

The safety assessments of the study include solicited and unsolicited, non-serious and serious adverse events (AEs) and medically attended AEs (MAAEs) reported in the study from the time of first dose of the vaccine. Participants were observed for 1-hour post vaccination to assess reactogenicity. Solicited local and systemic reactions were recorded for 7 consecutive days (Day 0-6), captured through subject diary after each vaccine dose. Solicited local AEs were pain, redness, swelling, itchy or warmth at injection site. Solicited systemic AEs were fever, headache, chills, Myalgia, arthralgia, fatigue, nausea, urticaria, rhinorrhoea, irritability, Hypotonic-hyporesponsive episodes, Somnolence, seizure and acute allergic reaction.

Unsolicited local and systemic adverse events (AEs) were recorded during the post-vaccination follow up period until 28 days after each dose. Serious adverse events (SAEs), medically attended adverse events (MAAEs) and adverse events of special interest (AESIs) if any, were collected during the entire study duration. Local and systemic reactions were scored by severity (mild, moderate, severe and life threatening) and the erythema and swelling or induration by the maximum diameter per day. Relatedness of study vaccine was also assessed for all reported AEs.

### Immunogenicity Analysis

Sera samples were collected from all the subjects in the immunogenicity cohort at Day-0 (pre-vaccination) and at Day-42 (14 days after second vaccine dose) time points. Following measurements were conducted to assess humoral immune response

1. Anti-RBD IgG concentration by ELISA, conducted at Dang’s Lab, India. The antibody concentrations were reported in ELISA Units/mL for each subject and Geometric Mean Concentrations were calculated for both time-points for both cohorts. % Seroconversion was also calculated at Day-42 timepoint for both cohorts
2. Neutralizing Antibody Titers (nAb titers) conducted at THSTI, India.^7^ The testing was conducted against Wild-Type SARS-COV-2 strains using Micro Neutralization Assay (MNA) in a BSL-3 facility at Translational Health Science and Technology Institute (THSTI), India. For prototype Wuhan Strain, the Victoria isolate from Australia was used while the Delta strain used in the assay was isolated in India.
3. Cellular immune response was assessed by ELISPOT method conducted at THSTI, India. Whole blood samples were collected post two-dose vaccination and PBMC’s were isolated and stored frozen. The PBMC’s were subsequently stimulated with various stimulants; SARS-COV-2 RBD peptides for specific response, DMSO for non-specific response and PHA for assay validity criteria. Post-stimulation, the number of PBMC’s that secrete cytokine Interferon-gamma were identified and quantified by ELISPOT technique and the Spot Forming Units (SFU’s) per million PBMC’s were calculated for each subject sample. Additional information in Supplementary section.

### Statistical Analyses

For the purposes of analysis, recruited subjects were further identified as total vaccinated cohort (TVC) and the according to protocol (ATP) cohort. All the demographic and primary safety analyses have been based on TVC population, defined as subjects who entered into the study and have received at least one single intramuscular dose of study vaccination.

ATP population is defined as population, who have blood samples available for immunogenicity analysis at all protocol specified time points from both CORBEVAX™ and COVISHIELD™ vaccinated cohorts. This has been the primary analysis population for immunogenicity assessment. The Geometric Mean Titers (nAb) were calculated post-vaccination against both Wuhan and Delta strains and then the ratio of the GMT’s for CORBEVAX™ to COVISHIELD™ cohort. Variances for each cohort were calculated from Log_10_ converted nAb titer values for each subject. Then the lower bound (LB) of the 95% confidence interval (CI) for the ratio of GMT’s were calculated via standard statistical methods. Superiority was concluded, if the lower limit of the one-sided 95% confidence interval (CI) for the ratio of two GMT’s) is >1.0. Non-inferiority to be inferred, if the lower limit of the one-sided 95% confidence interval (CI) for the ratio of two GMT’s is ≥0·67.

All data were summarized descriptively and data listings were based on all subjects enrolled in the study. By default, descriptive statistics for quantitative measurements included the number of subjects (n), mean, standard deviation (SD), minimum, median (IQR) and maximum. Safety data were summarised by System Organ Class and Preferred term. Serious adverse events, related adverse events, adverse events leading to death or withdrawal, solicited adverse events, medically attended adverse events and adverse events of special interest were summarised separately. In addition, adverse events were also summarized by severity. All analyses were conducted using SAS^®^ Version 9·4 or higher.

### Role of the funding source

BIRAC-a division of the Department of Biotechnology, Govt of India provided partial funding for the execution of trials. CEPI provided support for nAb titer testing in terms of reagents. Funding sources were not involved in the study conduct, data analysis/interpretation or writing the manuscript.

## RESULTS

### Participants

A total of 6485 subjects were screened and 2140 subjects were randomized in the study. One subject did not received vaccination randomized into immunogenicity arm (subject choice). So, in total 2139 subjects were enrolled into either immunogenicity arm (n=639) or safety arm (n=1500). Immunogenicity arm has two groups, receiving two doses of CORBEVAX™ vaccine (n=319) or COVISHIELD™ vaccine (n=320). Total vaccinated subjects with CORBEVAX™ vaccine including subjects from both immunogenicity and safety arm were n=1819. All subjects in the study were of Indian origin (100%). Demographic characteristics were comparable between subjects vaccinated with CORBEVAX™ or COVISHIELD™. Median age of CORBEVAX™ vaccinated cohort was 34 (IQR (Q1:Q3), 27·0:43·0) and COVISHIELD™ vaccinated cohort was 32 (IQR (Q1:Q3), 26·0:41·0) in years. Male: female ratio was 1283(70·5%): 536(29·5%) and 242(75·6%): 78(24·4%) in CORBEVAX™ or COVISHIELD™ vaccinated cohorts respectively. Other demographic and baseline characteristics of vaccinated (CORBEVAX™ or COVISHIELD™) were presented in table 1.

**Table 1:** Demographic characteristics of study participants

| Parameter | Statistic/Category<br>n (%) | CORBEVAX™<br>(N=1819) | COVISHIELD™<br>(N=320) |
| --- | --- | --- | --- |
| Age (years) | N | 1819 | 320 |
|  | Mean | 36.2 | 34.8 |
|  | Median | 34.0 | 32.0 |
|  | Range (Min: Max) | (18.0:79.0) | (18.0:77.0) |
|  | IQR (Q1:Q3) | (27.0:43.0) | (26.0:41.0) |
| Age Group | 18-44 | 1427 (78.5%) | 263 (82.2%) |
|  | 45-59 | 291 (16.0%) | 38 (11.9%) |
|  | 60-80 | 101 (5.6%) | 19 (5.9%) |
| Nationality | Indian | 1819 | 320 |
| Gender | Male | 1283 (70.5%) | 242(75.6%) |
|  | Female | 536 (29.5%) | 78(24.4%) |
| Height (cm) | N | 1819 | 320 |
|  | Mean | 164.0 | 163.3 |
|  | Median | 165.0 | 164.0 |
|  | Range (Min:Max) | (135.0:190.0) | (136.0:185.0) |
|  | IQR (Q1:Q3) | (158.0:170.0) | (158.0:169.0) |
| Weight (kgs) | N | 1819 | 320 |
|  | Mean | 64.6 | 63.6 |
|  | Median | 65.3 | 64.5 |
|  | Range (Min:Max) | (32.3:103.0) | (36.0:89.9) |
|  | IQR (Q1:Q3) | (58.4:70.5) | (55.6:72.0) |
| BMI | N | 1819 | 320 |
|  | Mean | 24.0 | 23.9 |
|  | Median | 23.7 | 23.8 |
|  | Range (Min:Max) | (12.5:42.3) | (15.0:40.6) |
|  | IQR (Q1:Q3) | (21.8:25.6) | (21.1:25.8) |

### Safety Findings

Safety data was presented for subjects enrolled into immunogenicity arm (n=639) including CORBEVAX™ vaccinated cohort (n=319) and COVISHIELD™ vaccinated cohort (n=320) and a safety cohort (n=1500) exclusively enrolled for safety assessment of CORBEVAX™ vaccine.

### Safety assessment of Immunogenicity group

Out of total 639 enrolled subjects, 68/319 (21·3%) subjects reported 84 events and 136/320 (42·5%) subjects reported 184 events in CORBEVAX™ and COVISHIELD™ arms respectively. CORBEVAX™ appeared to cause fewer local and systemic adverse reactions/events. The safety profile of CORBEVAX™ was comparable to the comparator vaccine COVISHIELD™ in terms of overall AE rates, related AE rates and medically attended AEs. All the reported adverse events were mild to moderate in their intensity and most of the reported adverse events were related to the study vaccine. Summary of AEs occurred in immunogenicity cohorts by system organ class (SOC) and preferred term (PT), severity grade and causality is listed in Table 2. Summary of local and systemic AEs by SOC and PT in immunogenicity arm and safety arm were reported as supplementary table 1 and supplementary table 2 respectively.

**Table 2:**
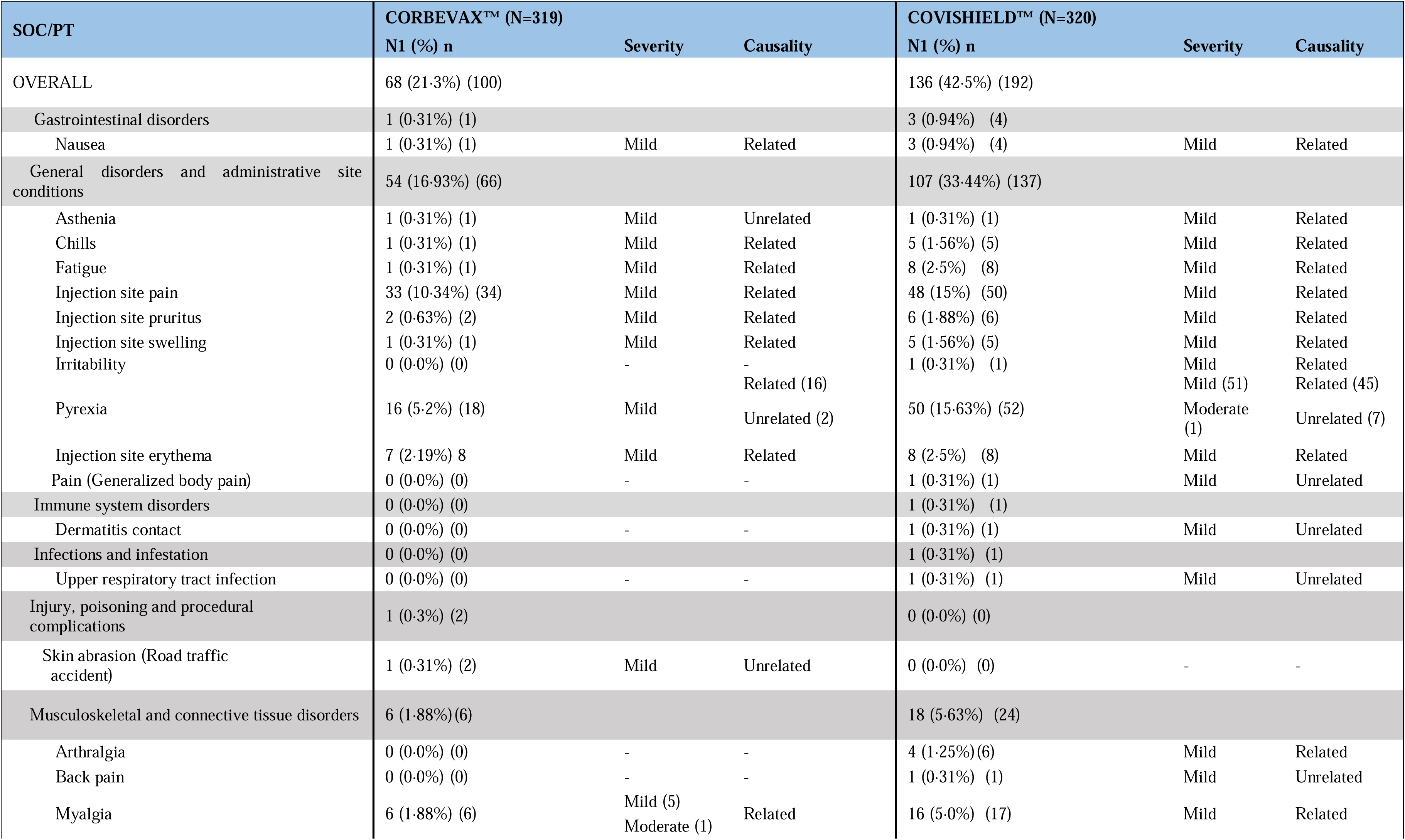
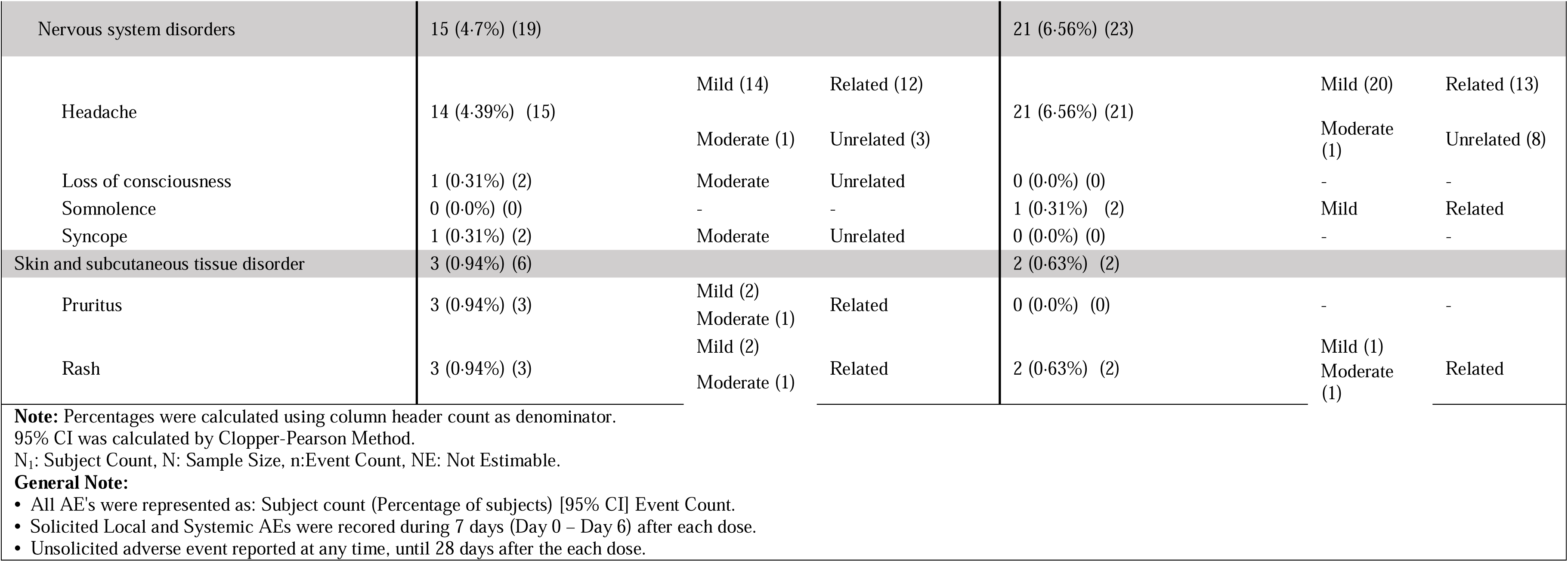
Summary of AEs by SOC and PT– Immunogenicity group (N=639); CORBEVAX™ Vs COVISHIELD™ vaccinated cohort

### Safety assessment of safety group

Out of total 1500 enrolled subjects, 361/1500 (24·1%) subjects reported 1221 events. The most commonly reported adverse events were Injection site pain [285 AEs in 267 (17·8%) subjects], Pyrexia [192 AEs in 184 (12·3%) subjects], Myalgia [158 AEs in 156 (10·4%) subjects], Headache [119 AEs in 115 (7·7%) subjects] and Fatigue [112 AEs in 109 (7·3%) subjects]. All the reported adverse events were mild to moderate in their intensity and most of the reported adverse events were related to the study vaccine (Table 3). Two serious AEs were reported in the safety group, which was of grade-3 severity and were diagnosed to be Dengue fever and Femur fracture. Causality of the events Dengue fever and femur fracture with the study vaccine (CORBEVAX™) is considered as not related by Principal Investigator and sponsor. There were no adverse events reported in the first 60 minutes’ post vaccination and no deaths reported in the study.

**Table 3:**
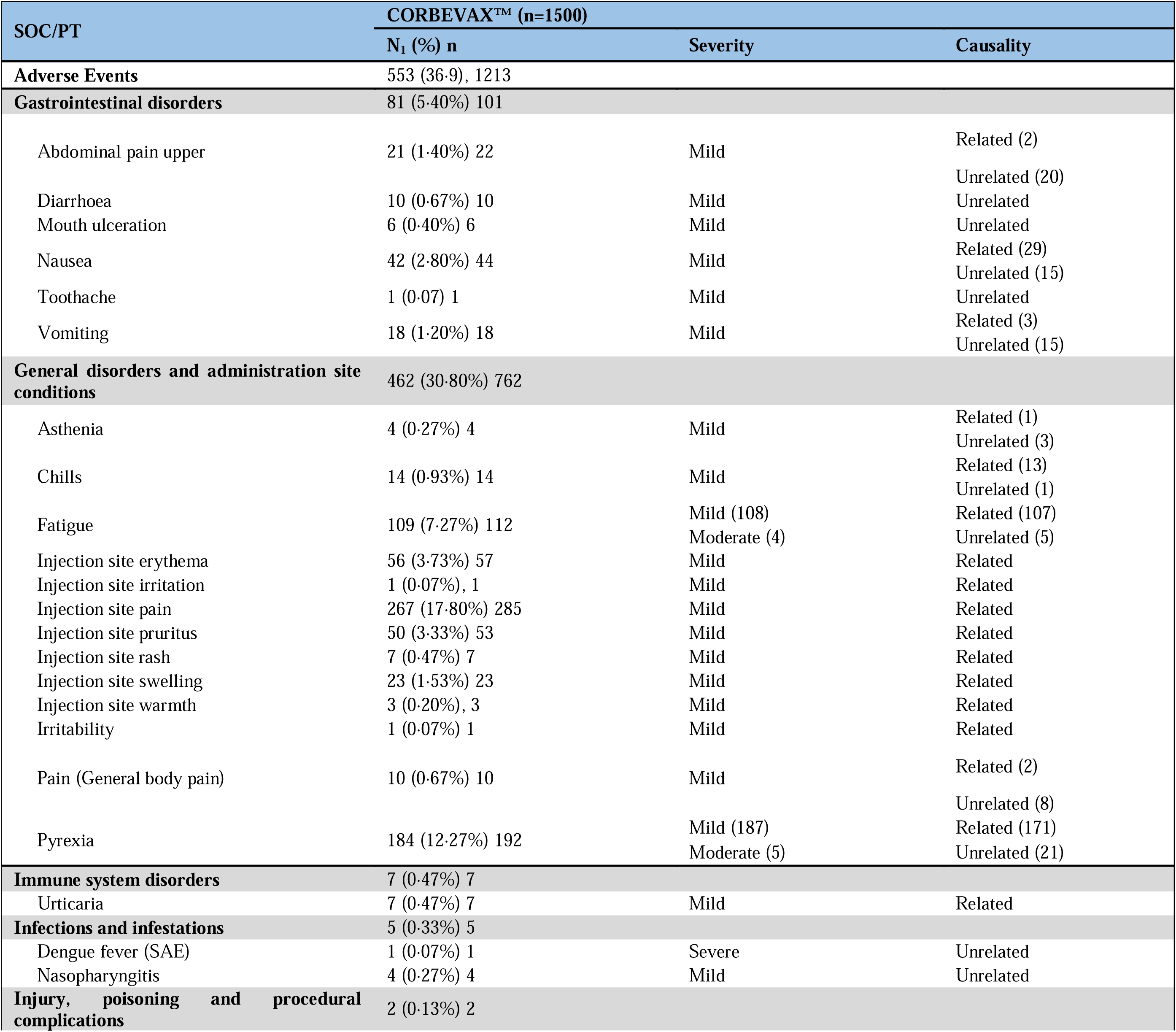
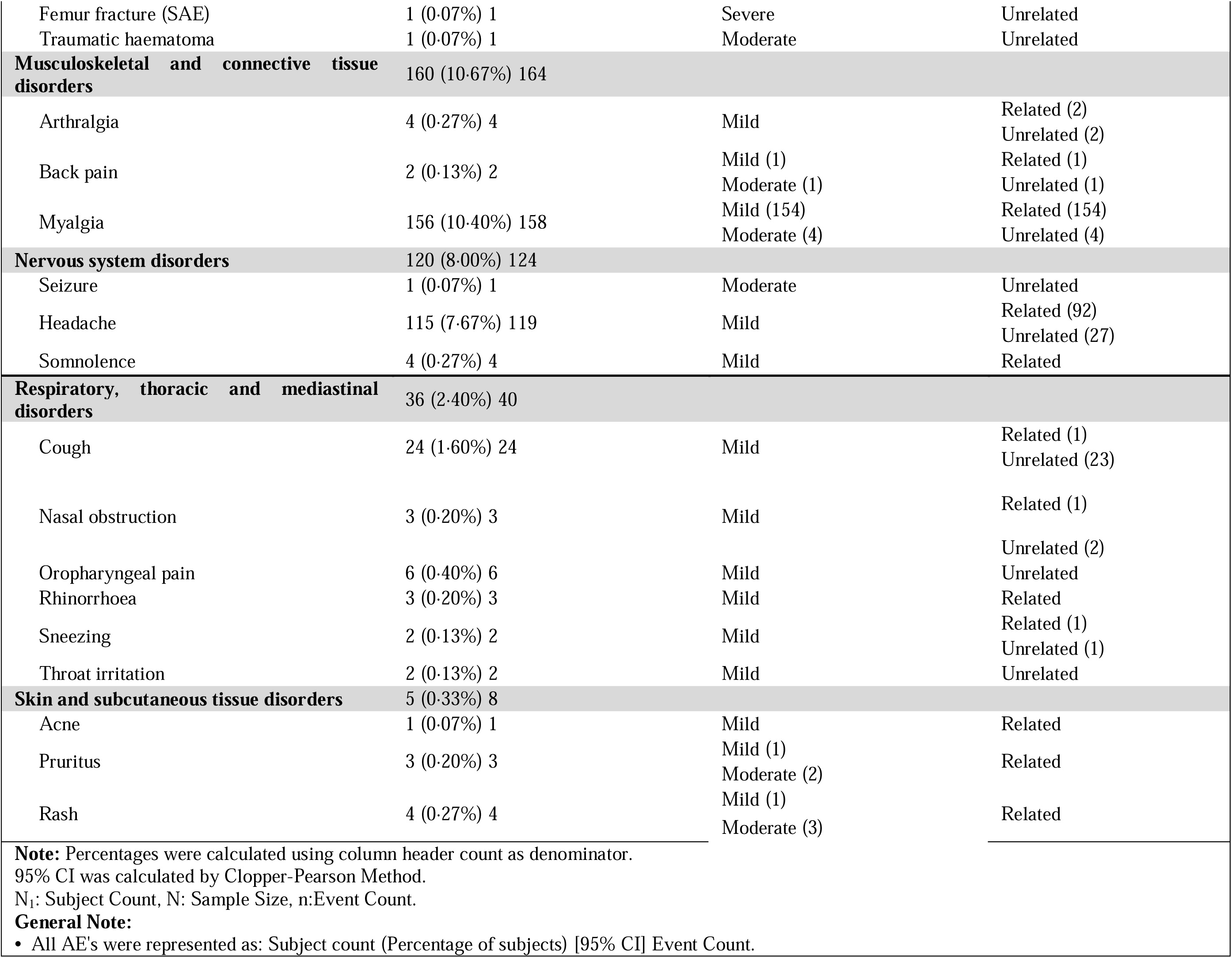
Summary of AEs by SOC and PT in subjects vaccinated with CORBEVAX™ vaccine

No marked changes overtime, were noted in the vital signs recorded. The AEs observed and physical examination results did not indicate any safety issues of concern. Majority of adverse events are mild to moderate in intensity and no AESI were reported in the study. Summary of AEs occurred in safety cohort by system organ class (SOC) and preferred term (PT), severity grade and causality is listed in Table 3.

Summary of local and systemic AEs by SOC and PT occurred in immunogenicity cohorts and in safety cohort are listed as supplementary table 1 and 2 respectively. Most of the systemic events resolved within 1 to 2 days. Most cases of fever resolved with antipyretic medications in 1 to 2 days. For fever occurring beyond 7^th^ day of each dose of vaccination, an RTPCR test for Covid-19 infection was done. None of them were positive for Covid-19 infection.

### Immunogenicity Findings

Humoral and cellular immune responses were evaluated from immunogenicity arm (n=639) of the study aimed to test immunogenic superiority of CORBEVAX™ vaccine (n=319) compared to COVISHIELD™ vaccine (n=320). Paired anti-RBD IgG concentration data at day 0 and day 42 were available in 304 subjects of CORBEVAX™ cohort and in 307 subjects of COVISHIELD™ cohort. Anti-RBD IgG concentrations (GMCs) increased significantly in both CORBEVAX™ and COVISHIELD™ vaccinated groups after the administration of two doses of vaccine compared to baseline (CORBEVAX™: 1439 EU/ml at day 0 Vs 24478 EU/ml at day 42; COVISHIELD™ : 1503 EU/ml at day 0 Vs 16203 EU/ml at day 42). However, the total antibody response against the RBD antigen is significantly higher in CORBEVAX™ cohort as compared to COVISHIELD™ cohort (24478 EU/ml Vs 16203 EU/ml at day 42) (Figure 2). Percent Seroconversion (SCR) was also calculated from the ratio of anti-RBD IgG concentration at Day-42-time point to Day-0 time-point i.e. post vs pre-vaccination. SCR was 91% in Corbavax vaccinated cohort and 88% in COVISHIELD™ vaccinated cohort.

**Figure 1:**
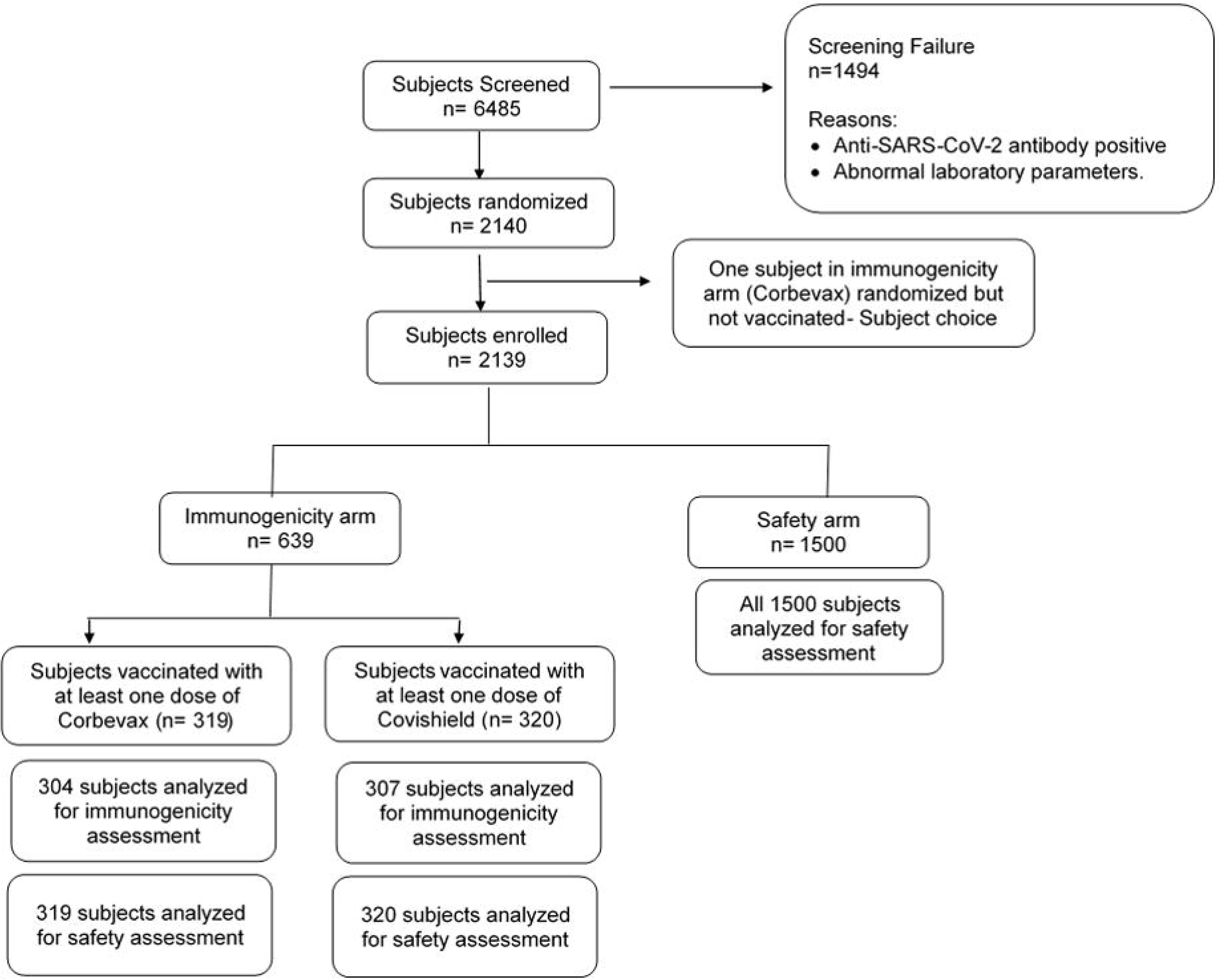
Study participants disposition.

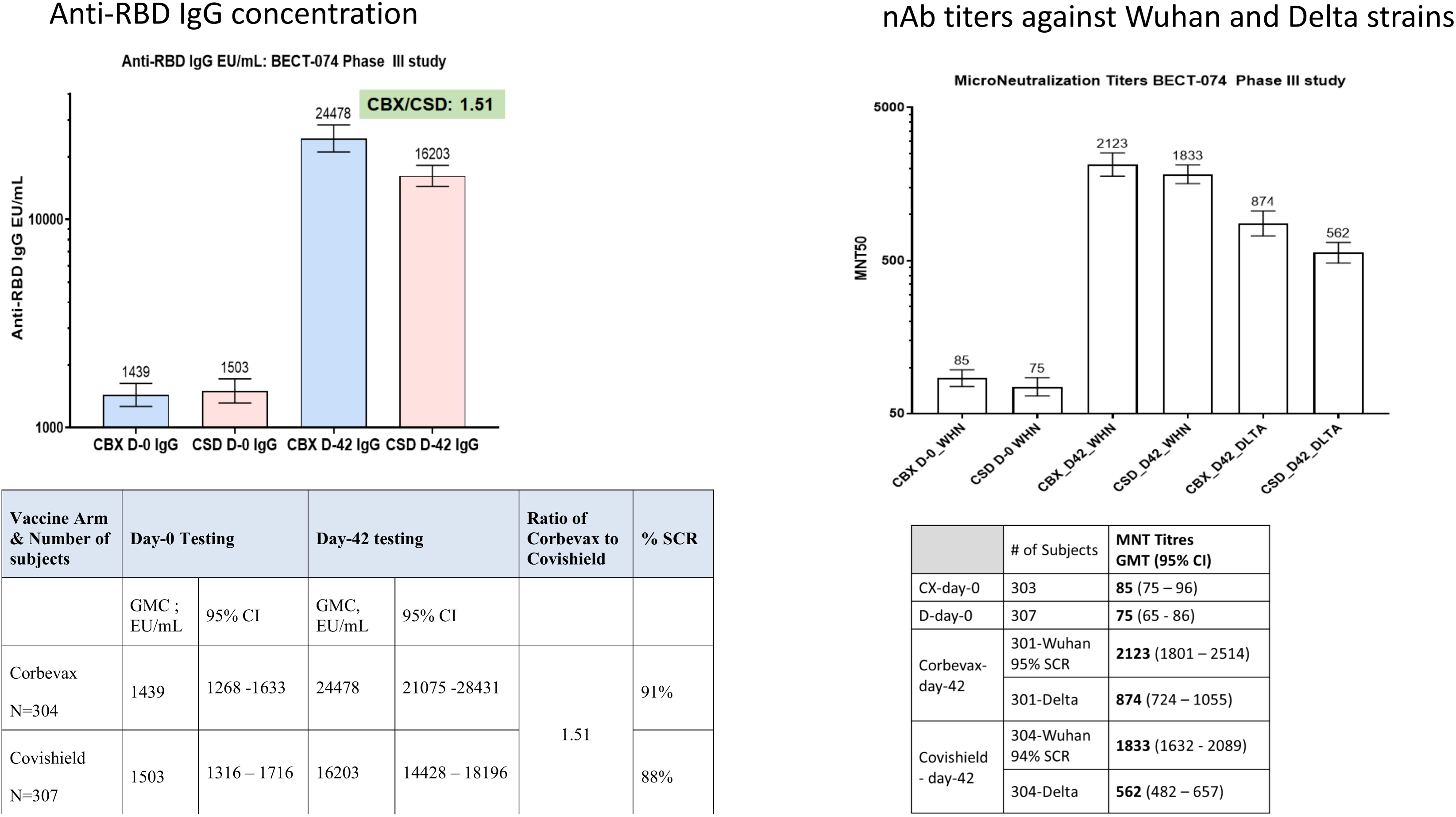

Neutralizing antibody (nAb) titers (GMTs) were assessed against the Wuhan and the Delta strain at baseline and day 42 in both CORBEVAX™ (n=303) and COVISHIELD™ (n=307) vaccinated cohorts. GMTs based on MNT50 when assessed against Ancestral strain were 85 (95% CI 75-96) and 75 (95% CI 65-86) at baseline and increased significantly at day 42 to 2123 (95% CI 1801 -2514) and 1833 (95% CI 1632 – 2089) in CORBEVAX™ and COVISHIELD™ cohorts respectively. GMTs based on MNT50 when assessed against delta strain were also significantly higher in CORBEVAX™ cohort (874; 95% CI 724-1055) as compared to COVISHIELD™ cohort (562; 95% CI 482 – 657). The CORBEVAX™ to COVISHIELD™ GMT ratios for Day-42 time-point were 1·15 and 1·56 respectively against Ancestral and Delta strains of SARS-COV-2 respectively. Using standard statistical techniques, the lower limit of the 95% confidence interval was determined as 1·02 and 1·27 for the GMT ratios against Ancestral and Delta strains respectively. Taken together, at day 42 (14 days after second vaccine dose) neutralizing antibody titers post-CORBEVAX™ vaccination is superior to COVISHIELD™ against both the Wuhan strain and Delta strains (Figure 2). CORBEVAX™ nAb GMT of 522 International Units/mL against Ancestral strain is indicative of vaccine effectiveness of >90% for prevention of symptomatic infections based on the Correlates of Protection assessment performed during Moderna and Astra-Zeneca vaccine Phase III studies. In a vaccine-effectiveness- (VE) trial conducted during the Delta-wave in India, THSTI observed COVISHIELD™ VE of 63% against symptomatic infection from Delta strain. Prior publication from Khoury et al^8^ estimated increase in VE from about 60% to 80% corresponding to approximately 50% increase in nAb titers. Thus, based on these observations, we can infer CORBEVAX™ to provide Vaccine Effectiveness of >80% against the dominant Delta strain.

Figure 3 shows the comparison of cellular responses in terms of ELISPOT data observed in a randomly selected sub-set of subjects in CORBEVAX™ and COVISHIELD™ cohorts. The CORBEVAX™ cohort had higher Interferon-gamma secreting PBMC’s post stimulation with SARS-COV-2 RBD peptides than COVISHIELD™ cohort in terms of average SFU’s and median SFU’s as summarized in Table 4.

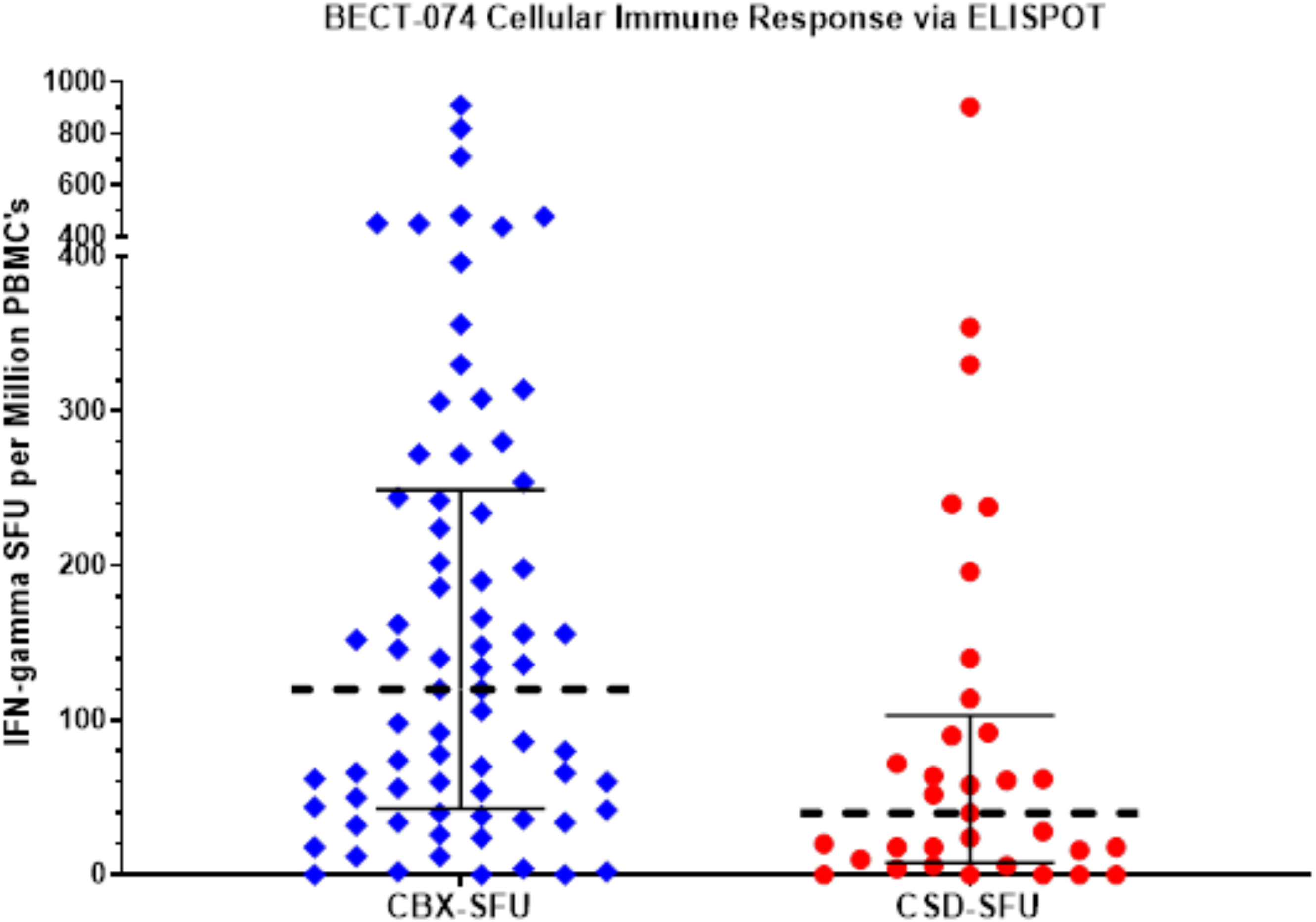

**Table 4:** Summary of interferon-gamma secreting PBMC’s as SFU’s per million PBMC’s in CORBEVAX™ and COVISHIELD™ cohorts

|  | Median (IQR) | Average |
| --- | --- | --- |
| CORBEVAX™ (N=73) | 120 (43-249) | 176 |
| COVISHIELD™ (N=33) | 40 (8 – 103) | 99 |

## DISCUSSION

In this trial, immunogenicity of a novel sub-unit vaccine for Covid-19 vaccine CORBEVAX™ was studied for safety and immunogenicity against RBD domain of SARS-CoV-2. Results indicated that CORBEVAX™ is a safe and well tolerated with no vaccine related serious adverse events, MAAEs or AESI when administered to adult individuals of Indian origin. This safety profile of CORBEVAX™ is comparable with that of another sub-unit vaccine Covavax.^9^ High immune responses in terms of anti-RBD IgG specific binding and protective antibodies were observed after second dose of vaccination. Here we also report, immunogenic superiority of CORBEVAX™ vaccine over COVISHIELD™, an adenoviral vector-based vaccine which is licensed in multiple countries, in terms of higher GMT’s of neutralizing antibodies against both the SARS-COV-2 Ancestral strain and the Delta strain.

In the present study, there were two SAEs reported which were unrelated to study vaccine: Dengue and a hip injury from a fall. In the immunogenicity arm of the study, only the subjects who were seronegative to SARS-CoV-2 were recruited, while in safety arm, seropositive subjects were also recruited because of rapidly increasing daily cases of COVID-19 in India (second wave), it was difficult to enroll seronegative subjects. Irrespective of the subjects being seronegative or seropositive at baseline, CORBEVAX™ vaccine was well tolerated by both the groups.

To establish relative immunogenicity of novel CORBEVAX™, we compared anti-RBD IgG antibody concentrations and neutralizing antibody titers in individuals receiving CORBEVAX™ or COVISHIELD™. While both CORBEVAX™ and COVISHIELD™ induced marked anti-RBD IgG Abs and neutralizing antibodies against Ancestral and Delta strains, responses and IFN-gamma cellular immune response induced by CORBEVAX™ were superior to that of COVISHIELD™. The nAb titers observed post CORBEVAX™ vaccination are indicative of very high vaccine effectiveness based on prior experience from efficacy studies performed for Moderna-mRNA1273 and AstraZeneca-AZD1222 vaccines.

### Study limitations

This study has several limitations like efficacy of the vaccine against Covid-19 infection was not studied and long-term safety was not established as interim results are available only until day 56. However, it is worth noting that in a small set of patients (n=360) safety was established until 12 months and significantly higher neutralizing antibody titres (nAbs) persisted at least 6 months after second dose of the vaccination when compared to human convalescent serum (HCS).^5^

Overall, we conclude that CORBEVAX™ is safe, well tolerated and elicited excellent antibody and cellular immune responses that can offer significant protection against symptomatic infection from SARS-CoV-2 virus.

## Data Availability

Additional study data which is not part of the manuscript can be made available upon request and addressed to the corresponding author Dr. Subhash Thuluva at his.

## Authors’ Contribution

ST and VP conceptualized the study and edited the manuscript for intellectual content. ST, SG, VY, RM and KT curated, accessed and verified the data and helped in interim report generation. VP, MK, SKM, SA, ASJ, GM and NG led the immunogenicity experiments. GM and NG contributed in performing and analyzing neutralizing antibody assays. AB, AZ and AA contributed in performing the ELISpot testing for cellular immune response assessment. CS and VRA were the key contributors of study conduct. ST was responsible for overall supervision of the project. All authors contributed to data interpretation, reviewing, and editing this manuscript.

## Acknowledgements

We are thankful to all the study participants, the principal investigators, and the study staff at all the clinical sites. In addition, we are thankful to the team at Dang’s Lab, New Delhi, led by Dr. Leena Chatterjee, Dr Arjun Dang, Mr Dinesh Kuma and Mr. Shakeeb Mohammad, performed the functions of study sample coordination (receipt, accessioning, aliquoting, labeling, storage, and dispatch) as well as conducted testing of all the ELISA assays (anti-RBD and cytokines) for all the samples. The study was funded by grants from BIRAC-a division of the Department of Biotechnology, Govt of India, and by the Coalition for Epidemic Preparedness Innovations. Dr. Maria Bottazzi and Dr. Peter Hotez, and their scientific team at the Center for Vaccine Development at Baylor College of Medicine/Texas Children’s Hospital, created and produced the recombinant Pichia Pastoris strain expressing the RBD protein. Dynavax, Inc supplied the adjuvant CpG1018 used in the CORBEVAX™ formulations. The clinical assay development team led by Dr. Arun Kumar at CEPI helped with neutralizing antibody titer assays in the supply of reagents and establishing assay consistency across multiple laboratories. Authors would like to thank Mr. Kamal Thammireddy for his valuable support in the study conduct. Development of this vaccine candidate would not have been possible without the efforts of manufacturing, quality control, quality assurance and regulatory teams from Biological E. The authors would like to thank Scientific Advisory Board (SAB) and management of Biological E Limited for their support and valuable guidance. All authors wish to express their appreciation and gratitude for all front-line healthcare workers.

## Declaration of interests

ST, VP, KT, SG, VY, RM, PVS, MK, SKM, SA and ASJ are employees of Biological E Limited and they don’t have any incentives or stock options. All other participating authors declare no competing interests.

---

**Supplementary Table 1:**
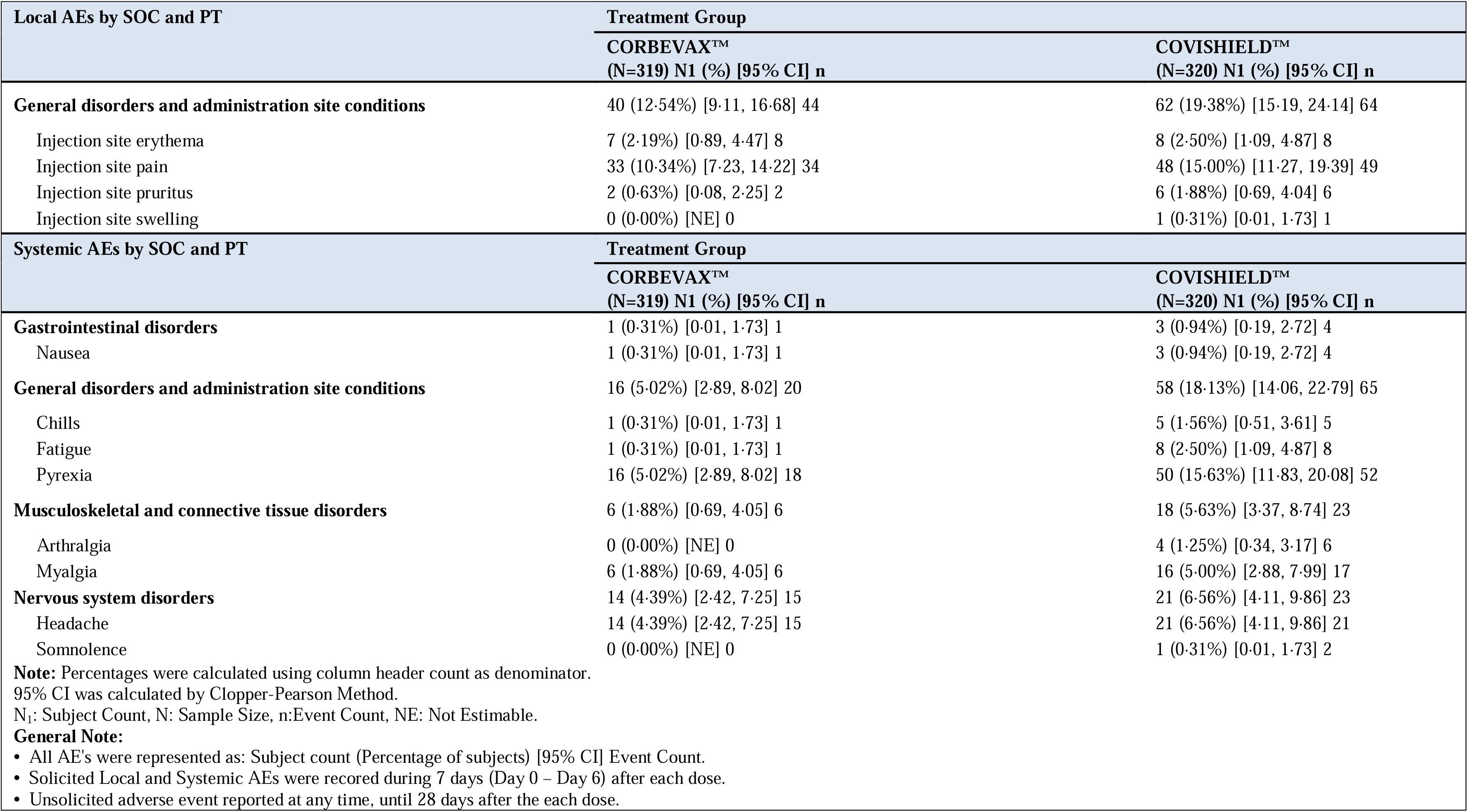
Summary of local and Systemic AEs by SOC and PT – Immunogenicity group (n=639)

| Local AEs by SOC and PT | Treatment Group |  |
| --- | --- | --- |
|  | CORBEVAX™<br>(N=319) N1 (%) [95% CI] n | COVISHIELD™<br>(N=320) N1 (%) [95% CI] n |
| **General disorders and administration site conditions** | 40 (12.54%) [9.11, 16.68] 44 | 62 (19.38%) [15.19, 24.14] 64 |
| Injection site erythema | 7 (2.19%) [0.89, 4.47] 8 | 8 (2.50%) [1.09, 4.87] 8 |
| Injection site pain | 33 (10.34%) [7.23, 14.22] 34 | 48 (15.00%) [11.27, 19.39] 49 |
| Injection site pruritus | 2 (0.63%) [0.08, 2.25] 2 | 6 (1.88%) [0.69, 4.04] 6 |
| Injection site swelling | 0 (0.00%) [NE] 0 | 1 (0.31%) [0.01, 1.73] 1 |
| Systemic AEs by SOC and PT | Treatment Group |  |
|  | CORBEVAX™<br>(N=319) N1 (%) [95% CI] n | COVISHIELD™<br>(N=320) N1 (%) [95% CI] n |
| **Gastrointestinal disorders** | 1 (0.31%) [0.01, 1.73] 1 | 3 (0.94%) [0.19, 2.72] 4 |
| Nausea | 1 (0.31%) [0.01, 1.73] 1 | 3 (0.94%) [0.19, 2.72] 4 |
| **General disorders and administration site conditions** | 16 (5.02%) [2.89, 8.02] 20 | 58 (18.13%) [14.06, 22.79] 65 |
| Chills | 1 (0.31%) [0.01, 1.73] 1 | 5 (1.56%) [0.51, 3.61] 5 |
| Fatigue | 1 (0.31%) [0.01, 1.73] 1 | 8 (2.50%) [1.09, 4.87] 8 |
| Pyrexia | 16 (5.02%) [2.89, 8.02] 18 | 50 (15.63%) [11.83, 20.08] 52 |
| **Musculoskeletal and connective tissue disorders** | 6 (1.88%) [0.69, 4.05] 6 | 18 (5.63%) [3.37, 8.74] 23 |
| Arthralgia | 0 (0.00%) [NE] 0 | 4 (1.25%) [0.34, 3.17] 6 |
| Myalgia | 6 (1.88%) [0.69, 4.05] 6 | 16 (5.00%) [2.88, 7.99] 17 |
| **Nervous system disorders** | 14 (4.39%) [2.42, 7.25] 15 | 21 (6.56%) [4.11, 9.86] 23 |
| Headache | 14 (4.39%) [2.42, 7.25] 15 | 21 (6.56%) [4.11, 9.86] 21 |
| Somnolence | 0 (0.00%) [NE] 0 | 1 (0.31%) [0.01, 1.73] 2 |
| **Note:** Percentages were calculated using column header count as denominator.<br>95% CI was calculated by Clopper-Pearson Method.<br>N<sub>1</sub>: Subject Count, N: Sample Size, n:Event Count, NE: Not Estimable.<br>**General Note:**<br>All AE's were represented as: Subject count (Percentage of subjects) [95% CI] Event Count.<br>Solicited Local and Systemic AEs were recored during 7 days (Day 0 – Day 6) after each dose.<br>Unsolicited adverse event reported at any time, until 28 days after the each dose. |  |  |

**Supplementary Table 2:**
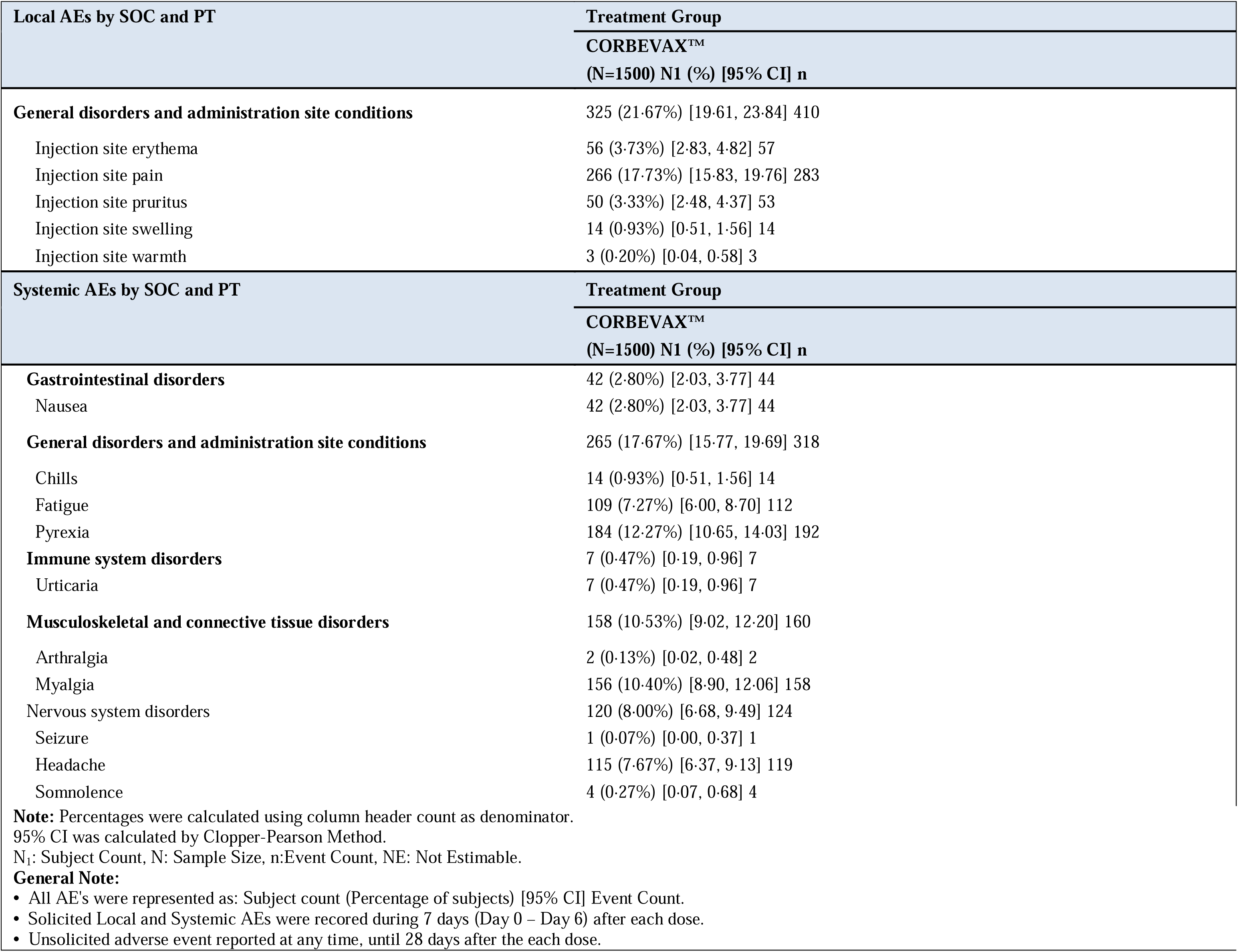
Summary of locl and Systemic AEs by SOC and PT – safety group (n=1500)

## Notes

### Competing Interest Statement

The authors have declared no competing interest.

### Clinical Trial

CTRI/2021/08/036074

### Author Declarations

The Investigational Review Board or Ethics Committee at each study site approved the protocol. Centre- 1: Prakhar Hospital, Kanpur, approved on30 Aug 21 Centre- 2: GTB Hospital, Delhi, approved on 14 Sep 21 Centre- 3: ESIC Medical College & Hospital, Faridabad, approved on 30 Aug 21 Centre- 4: Shubham Sudbhawana Hospital, Varanasi, approved on 04 Sep 21 Centre- 5: St.Theresas Hospital (STH), Hyderabad, approved on 30 Aug 21 Centre- 6: KLES Dr. Prabhakar Kore Hospital & Medical Research Centre, Belgavi, approved on 03 Sep 21 Centre- 7: AIG Hospital, Hyderabad, approved on14 Sep 21 Centre- 8: Belgavi Institute of Medical Sciences, Belagavi, approved on 13 Sep 21 Centre- 9: National Institute of Medical Sciences (NIMS), Jaipur, approved on 11 Sep 21 Centre- 10: Grant Medical College & Sir J.J Hospital, Mumbai, approved on 13 Sep 21 Centre- 11: JLN Medical College, Ajmer, approved on 21 Sep 21 Centre- 12: Christian Medical College & Hospital, Ludhiana, approved on 25 Sep 21 Centre- 13: Apex Hospital, Jaipur, approved on 09 Sep 21 Centre- 14: Medanta Institute of Education and Research, Gurgaon, approved on 09 Sep 21 Centre- 15: BAPS Pramukh Swami Hospital, Surat06 Sep 21 Centre- 16: Christian Medical College Vellore Association, Vellore, approved on 30 Sep 21 Centre- 17: JSS Hospital, Mysuru, approved on 16 Sep 21 Centre- 18: Mahatma Gandhi Institute of Medical Sciences (MGIMS), Wardha, approved on 04 Sep 21 Centre- 19: All India Institute of Medical Sciences (AIIMS), Patna, approved on 09 Sep 21 Centre- 20: Samvedna Hospital, Varanasi, approved on 11 Sep 21

## REFERENCES

1. Sanyaolu A, Okorie C, Marinkovic A, et al. Comorbidity and its Impact on Patients with COVID-19. SN Compr Clin Med. 2020;2:1069–76.

2. Krammer F. SARS-CoV-2 vaccines in development. Nature. 2020;586:516–527.

3. Chen WH, Pollet J, Strych U, et al. Yeast-expressed recombinant SARS-CoV-2 receptor binding domain RBD203-N1 as a COVID-19 protein vaccine candidate. Protein Expr Purif. 2022;190:106003.

4. Yang J, Wang W, Chen Z, et al. A vaccine targeting the RBD of the S protein of SARS-CoV-2 induces protective immunity. Nature 2020; 586(7830): 572–7.

5. Thuluva S, Paradkar V, et al. Selection of optimum formulation of RBD-based protein sub-unit covid19 vaccine (CORBEVAX) based on safety and immunogenicity in an open-label, randomized Phase-1 and 2 clinical studies medRxiv 2022.03.08.22271822.

6. LIAISON® SARS-CoV-2 S1/S2 IgG. The fully automated serology test for the detection of SARS-CoV-2 IgG Antibodies. Available from https://www.diasorin.com/sites/default/files/allegati/liaisonr_sars-cov-2_s1s2_igg_brochure.pdf.pdf. Last accessed on 17 March 2022.

7. Bewley KR, Coombes NS, Gagnon L, et al. Quantification of SARS-CoV-2 neutralizing antibody by wild-type plaque reduction neutralization, microneutralization and pseudotyped virus neutralization assays. Nat Protoc 2021;16:3114–40.

8. Khoury DS, Cromer D, Reynaldi A, et al. Neutralizing antibody levels are highly predictive of immune protection from symptomatic SARS-CoV-2 infection. Nat Med. 2021;27:1205–11.

9. Heath PT, Galiza EP, Baxter DN, et al; 2019nCoV-302 Study Group. Safety and Efficacy of NVX-CoV2373 Covid-19 Vaccine. N Engl J Med. 2021;385:1172–83.

