## Supplementary Information for "Immunogenic superiority and safety of Biological E’s CORBEVAX™ vaccine compared to COVISHIELD™ (ChAdOx1 nCoV-19) vaccine studied in a phase III, single blind, multicenter, randomized clinical trial"

**Supplementary Material**

**Inclusion Criteria**

Subjects were enrolled in the study based on the following inclusion criteria.

1. Subject is seronegative to anti-SARS-CoV-2 IgG antibody prior to randomisation either into Group-1 and Group-2
2. Subject is virologically seronegative to SARS-CoV-2 infection as confirmed by RT-PCR test prior to enrolment in all groups
3. Male or female subject between ≥ 18 to 80 years of age
4. Subject is willing to provide a written informed consent for voluntary participation in the study
5. Subject, in the opinion of the investigator, has ability to communicate and willingness to comply with the requirements of the protocol
6. Subject is seronegative to HIV 1 and 2, HBV and HCV infection prior to enrolment
7. Subject is considered of stable health as judged by the investigator, determined by medical history and physical examination
8. Female subject of childbearing potential must have a negative urine pregnancy test (UPT), and willingness to avoid becoming pregnant through use of an effective method of contraception or abstinence from the time of study enrolment until six weeks after the last dose of vaccination in the study
9. Male subject, who is sexually active, must agree to use double-barrier contraception (e.g. condom with spermicide) with his female partner during the study period. Male subject should also agree to avoid semen donation or providing semen for in-vitro fertilization during the study duration
10. Subject agrees not to participate in another clinical trial at any time during the total study period
11. Subject agrees to refrain from blood donation during the course of the study
12. Subject agrees to remain in the town where the study centre is located, for the entire duration of the study

**Exclusion Criteria**

Subjects were excluded from the study based on the following exclusion criteria:

1. History of vaccination with any investigational or approved vaccine against COVID-19 disease
2. Subject living in the same household as that of any active COVID-19 positive individual
3. History of receipt of any licensed vaccine within 1 month prior to screening, likely to impact on interpretation of the trial data (e.g., influenza vaccines);
4. Subjects with any clinically significant abnormal haematology and biochemical laboratory parameters tested at screening as judged by the investigator
5. Subjects with Body temperature of ≥100.4°F (>38.0°C) or symptoms of an acute illness at the time of screening or prior to vaccination
6. Pregnant women, nursing women or women of childbearing potential who are not actively avoiding pregnancy during the study
7. Subjects with known current or chronic history of any of the following conditions, likely to affect participation in the study
8. severe psychiatric conditions;
9. any bleeding disorder (e.g. factor deficiency, coagulopathy or platelet disorder);
10. allergic disease or reactions likely to be exacerbated by any component of the study vaccine (BE SARS-CoV-2 COVID-19 vaccine);
11. neurological illness, and any other serious chronic illness requiring hospital specialist supervision
12. Subjects requiring chronic administration (defined as more than 14 days in total) of immunosuppressant (e.g. corticosteroids, cytotoxic drugs or antimetabolites, etc.) or other immune-modifying drugs (e.g. interferons) during the period starting six months prior to the first vaccine dose including use of any blood products
13. For corticosteroids, this will mean prednisone ≥0.5 mg/kg/day, or equivalent
14. Inhaled and topical steroids are allowed
15. Receipt of prohibited concomitant medication that may jeopardize the safety of the participant or interpretation of the data
16. Any confirmed or suspected immunosuppressive or immunodeficient condition, based on medical history and physical examination (no laboratory testing required)
17. Any medical condition that in the judgment of the investigator would make study participation unsafe
18. Planned use of any investigational or non-registered product other than the study vaccine during the trial period or 3 months prior to enrolment
19. Current or planned participation in prophylactic drug trials for the duration of the study
20. Individuals who are part of the study team or close family members of individuals conducting the study

**Methodology**

As per the kit manufacturer, subjects with antibody concentrations below 12 Antibody Units/mL were designated as sero-negative and were selected in the trial (Diasorin kit). Health status assessed during the screening period was based on medical history and clinical laboratory findings, vital signs, and physical examination. All those who were part of any other clinical trial, with a history of vaccination with any investigational vaccine against Covid-19 disease, any other health issues, or were on immunosuppressants, immunodeficient conditions or sero-positive for SARS-COV-2 were excluded from the study.

**Procedure**

**Safety Assessments:**

The number and percentage of subjects with Adverse events (AEs) and severe adverse events (SAEs) were presented overall by system organ class (SOC) & preferred term (PT). The percentage of subjects with at least one local AE (solicited and unsolicited), with at least one systemic AE (solicited and unsolicited) and with any AE during the solicited follow-up period were tabulated with exact 95% confidence interval (CI). The same calculations were performed for symptoms rated as Grade 3 and above. Systemic and local tolerability, recorded in subject diaries, were summarized in a frequency table with percentages based on the number of observed values. Serious adverse events, related AEs, AEs leading to death or withdrawal, solicited AEs, and MAAEs were summarized separately.

All reported AEs during the entire study period, was summarized by calculating frequencies and were listed per subject including severity, relationship to the vaccine (causality) and action taken with the vaccine. All AEs were coded using the Medical Dictionary for Regulatory Activities (MedDRA) coding dictionary and concomitant medications were coded using the World Health Organization (WHO) Drug Dictionary.Summary of clinically significant increase in body temperature across study visits were tabulated and was summarized descriptively. Also mean change in the body temperature at each visit was presented exploratorily only if clinically significant. All SAEs and medically attended AEs reported during the study (start and up to end of the study) were listed and analysed for expectedness and causality.

**Immunogenicity Analysis**

Humoral immune responses were evaluated by following methods:

1. Anti-RBD Antibody response: Anti-RBD IgG concentration in the subject sera samples were measured at pre-vaccination (Day-0), post first dose (Day-28) and at multiple time-points post second dose (Day-42, Day-56, Day-208) using a validated enzyme linked immunosorbent assay (ELISA) method executed at Dang’s Lab, New Delhi, India. A monoclonal antibody, CR-3022 supplied by Lake Pharma Inc, CA, USA, that binds specifically to the RBD protein was used to generate standard curve of ELISA OD response vs. antibody concentrations. ELISA concentration equivalent to 1 ng/mL of CR-3022 antibody binding concentration was assigned concentration of 1 anti-RBD ELISA Unit/mL and thus anti-RBD IgG concentration in the sera samples were reported in EU/mL. The National Institute for Biological Standards and Control, UK plasma reference standard 20/130 was used as a positive control on all the plates with a control range of (**8151** to **15137** EU/mL) for validity consideration^1^. Geometric means were calculated for various subject cohorts at specific time-points and fold rise in anti-RBD concentrations for all time-points post start of vaccination were calculated in relation to the pre-vaccination concentrations and then geometric mean fold rise (GMFR) were calculated for each cohort.
2. SARS-COV-2 Virus Neutralization: SARS-COV-2 neutralizing antibody titers (nAb titers) were measured via Microneutralization Assay (MNA) using Wild-Type SARS-COV-2 strain (Victoria isolate 01/2020). The MNA was conducted at Translational Health Science and Technology Institute (THSTI), Faridabad, India; which is a participating laboratory in Coalition for Epidemic Preparedness Innovation (CEPI) - network. The nAb testing was conducted as per methods described previously^2^. Conversion factors have been established to enable conversion of the Neutralization Titer (NT_50_) values to WHO-International Standard (NIBSC-20/136) and report the (NT_50_ values in International Units/mL^3^. MNA values were divided by 4.064 obtain the titers in IU/mL when required for comparison. Geometric mean titers were calculated at scheduled time-points and fold Rise from the pre-vaccination values were calculated along with GMFR. Sera samples that did not demonstrate minimum 50% neutralization of the virus at the initial dilution i.e. the of the assay, titers were assigned as LLOQ/2. For key GMT/GMC values, 95% Confidence Intervals (95%CI) were also calculated. All the subject sera samples were tested for nAb titers using PNA method. The subjects that had PNA titers below the LLOQ were considered as sero-negative for analysis while subjects with PNA titers above LLOQ were considered as sero-positive for analysis. For samples that were below the level of quantitation (LLOQ), the titers were assigned as half of the LLOQ multiplied by the dilution factor. For samples that were above the level of quantitation (ULOQ), the titers were assigned as two times the ULOQ of the assay multiplied by the dilution factor of the sample. The LLOQ and ULOQ titers are 20 and 640 respectively. These samples were tested at different dilutions to obtain the titer information in subsequent testing. Seroconversion rate for anti-RBD IgG concentration and nAb titers were calculated as follows: For sero-negative subjects, ≥4‑fold rise in IgG or nAb titers was considered to be seroconverted while for sero-positive subjects, ≥2‑fold rise in IgG or nAb titers was considered to be seroconverted.
3. Cellular immune responses were assessed in a randomly selected subset of subjects in terms of Interferon-gamma secreting PBMC’s post stimulation with SARS-COV-2 RBD peptides to detect an antigen-specific T-cell immune response. This was done using the Interferon-gamma ELISpot assay. The enzyme-linked immune absorbent spot (ELISpot) relies on visualizing cytokine secretion by individual T cells following *in vitro* stimulation with antigen. This assay identifies biologically active, cytokine-secreting cells from isolated peripheral blood mononuclear cells (PBMC), at the single-cell level. It is a highly sensitive technique that detects the presence of IFN-γ-producing CD4+ and/or CD8+ T cells following their stimulation with specific antigens. The PBMC’s isolated from whole blood samples collected from subjects were resuspended in appropriate growth medium and from each subject sample was added to six wells (0.25 million PBMC’s per well) in MAbtech ELISpot plate. Three stimulants were added to the wells (each in two wells): SARS-COV-2 RBD peptide pool (procured from JPT, Berlin, Germany) for antigen-specific stimulation assessment, DMSO for non-specific stimulation assessment and PHA for assay validity assessment. After 20 hrs of incubation, the plate was washed with PBS and the ELISpot assay was performed according to the MAbtech ELISPOT assay kit’s manufacturer’s instructions. Briefly, the detection antibody was added to the wells at a 1µg/ml concentration, and the plate was incubated for 2 hours at room temperature. The plate was again washed, and Streptavidin-ALP was added to the wells and left for incubation for 1 hour at room temperature. This was followed by adding a filtered ready-to-use substrate solution for developing the spots until distinct spots emerged. The color development was stopped by washing the plate extensively with deionized water. The plate was then left to dry overnight, and the spots were quantified as Spot Forming Units (SFU’s) using an AID iSPOT reader on the next day. The antigen specific Spot Forming Units were calculated by subtracting the SFU’s from DMSO stimulation from the SFU’s observed post stimulation with SARS-COV-2 RBD peptides. The Interferon-gamma SFU’s were then reported in terms of million PBMC’s for each subject. Additional details of the assay are provided in the preprint version of the manuscript by Thiruvengadam et al^4^.

**List of study Investigators**

| **S. No.** | **Site**  **Code** | **Name of the Investigator** | **Institutional Affiliations and Site Locations** |
| --- | --- | --- | --- |
| 1. | 02 | Dr.J.S Khushwaha  M.B.B.S, MD  (General Medicine) | Prakhar Hospital, 8/219, Khalasi Line,  Arya Nagar, Kanpur 208002, Uttar  Pradesh, India |
| 2. | 03 | Dr.Shiva Narang M.B.B.S, MD (General Medicine) | GTB Hospital, Delhi,  Tahirpur Rd,  GTB Enclave, Dilshad Garden,  Delhi 110095, India |
| 3. | 05 | Dr.Anil Kumar Pandey M.B.B.S, MD (General Medicine) | ESIC Medical College & Hospital, Room No. 440, 4th Floor, NH-3 behind BK Hospital New Industrial Town, Faridabad-121001, Haryana, India |
| 4. | 06 | Dr.Indranil Basu M.B.B.S, MD (General Medicine) | Shubham Sudbhawana Hospital, B 31/80, 23B - Bhogabeer, Lanka, Varanasi, 221005, Uttar Pradesh, India |
| 5. | 07 | Dr.A.Venkateshwara Rao M.B.B.S, DNB | St.Theresas Hospital (STH), 1st Floor, Room No. 05, Erragadda Main Road, Czech Colony Sanath Nagar, Hyderabad 500038, Telangana, India |
| 6. | 09 | Dr.Madhav Prabhu M.B.B.S, MD (General Medicine | KLES Dr. Prabhakar Kore Hospital & Medical Research Centre, Department of Medicine, Nehru Nagar,Belgavi-590010, Karnataka, India |
| 7. | 10 | Dr.P.Naveen Chander Reddy M.B.B.S, MD (General Medicine | AIG Hospital, 4th floor, Plot No. 2/3/4/5, Survey No. 136/1, Mindspace Road, Gachibowli, Hyderabad--500032, Telanagana, India |
| 8. | 11 | Dr.Rachgonda G.Viveki M.B.B.S, MD (Community Medicine) | Belgavi Institute of Medical Sciences, Dr B R Ambedkar Rd, Sadashiv Nagar, Belagavi - 590019, Karnataka, India |
| 9. | 12 | Dr.Rajendra Dhar M.B.B.S, MD (General Medicine) | National Institute of Medical Sciences (NIMS), NH-11C, Delhi - Jaipur Expressway, Shobha Nagar, Jaipur 303121, Rajasthan, India |
| 10. | 16 | Dr.Khobragade Akash Ashok Kumar M.B.B.S, MD (General Medicine) | Grant Medical College & Sir J.J Hospital, J J Marg, Nagpada, Mumbai Central, Mumbai 400008, Maharashtra, India |
| 11. | 18 | Dr.Veer Bhadur Singh M.B.B.S, MD (General Medicine) | JLN Medical College , Kala Bagh, Ajmer 305001, Rajasthan, India |
| 12. | 19 | Dr.Clarence.J Samuel M.B.B.S, MD (Community Medicine) | Christian Medical College & Hospital, Brown Rd, CMC Campus, Ludhiana - 141008, Punjab, India |
| 13. | 20 | Dr.Vipul Khandelwal M.B.B.S, MD (Internal Medicine) | Apex Hospital, SP 4& 6 MIA Malviya Nagar near apex circle, Jaipur-302017, Rajasthan, India |
| 14. | 21 | Dr. Sushila Kataria M.B.B.S, MD (General Medicine) | Medanta Institute of Education and Research, Sector 38, CH Baktawar Singh Rd, Medicity, Islampur Colony, Sector 38, Gurgaon - 122018, Haryana |
| 15. | 23 | Dr. Parshottam Koradia M.B.B.S, MD (Internal Medicine) | BAPS Pramukh Swami Hospital, Shri Pramukh Swami Maharaj Marg, Adajanchar rasta. Adajan. Surat- 395 009, Gujarat, India |
| 16. | 24 | Dr. Winsley Rose M.B.B.S, MD (Paediatrics) | Christian Medical College Vellore Association, IDA Scudder Rd, Vellore 632004, Tamil Nadu, India |
| 17. | 25 | Dr Shilpa Avarebeel M.B.B.S, DNB (General Medicine) | JSS Hospital, Mahatma Gandhi Road, Fort Mohalla, Mysuru-570004, Karnataka, India |
| 18. | 26 | Dr. Bishan Swarup Garg M.B.B.S, MD (Community Medicine) | Mahatma Gandhi Institute of Medical Sciences (MGIMS), Sewagram, Wardha 442102, Maharashtra, India |
| 19. | 28 | Dr.Chandramani Singh M.B.B.S, MD (Community Medicine) | All India Institute of Medical Sciences (AIIMS), Room No. 17 Department of Community & Family Medicine, Aurangabad Road Phulwari Sharif, Patna 801507, Bihar, India |
| 20. | 29 | Dr. Monica Gupta M.B.B.S, MD (General Medicine) | Samvedna Hospital, B-27/88-G, New Colony, Varanasi-221005, Uttar Pradesh, India |

1. **Institutional Ethics Committees (IECs) and approvals**

| **Centre Code (Name)** | **Date of EC approval** |
| --- | --- |
| **Centre- 1:** Prakhar Hospital, Kanpur | 30 Aug 21 |
| **Centre- 2:** GTB Hospital, Delhi | 14 Sep 21 |
| **Centre- 3:** ESIC Medical College & Hospital, Faridabad | 30 Aug 21 |
| **Centre- 4:** Shubham Sudbhawana Hospital, Varanasi | 04 Sep 21 |
| **Centre- 5:** St.Theresas Hospital (STH), Hyderabad | 30 Aug 21 |
| **Centre- 6:** KLES Dr. Prabhakar Kore Hospital & Medical Research Centre, Belgavi | 03 Sep 21 |
| **Centre- 7:** AIG Hospital, Hyderabad | 14 Sep 21 |
| **Centre- 8:** Belgavi Institute of Medical Sciences, Belagavi | 13 Sep 21 |
| **Centre- 9:** National Institute of Medical Sciences (NIMS), Jaipur | 11 Sep 21 |
| **Centre- 10:** Grant Medical College & Sir J.J Hospital, Mumbai | 13 Sep 21 |
| **Centre- 11:** JLN Medical College, Ajmer | 21 Sep 21 |
| **Centre- 12:** Christian Medical College & Hospital, Ludhiana | 25 Sep 21 |
| **Centre- 13:** Apex Hospital, Jaipur | 09 Sep 21 |
| **Centre- 14:** Medanta Institute of Education and Research, Gurgaon | 09 Sep 21 |
| **Centre- 15:** BAPS Pramukh Swami Hospital, Surat | 06 Sep 21 |
| **Centre- 16:** Christian Medical College Vellore Association, Vellore | 30 Sep 21 |
| **Centre- 17:** JSS Hospital, Mysuru | 16 Sep 21 |
| **Centre- 18:** Mahatma Gandhi Institute of Medical Sciences (MGIMS), Wardha | 04 Sep 21 |
| **Centre- 19:** All India Institute of Medical Sciences (AIIMS), Patna | 09 Sep 21 |
| **Centre- 20:** Samvedna Hospital, Varanasi | 11 Sep 21 |

**References**.

1.	NIBSC. Data Sheet -Research reagent for anti-SARS-CoV-2 Ab NIBSC code 20/130.

2.	Bewley KR, Coombes NS, Gagnon L, et al. Quantification of SARS-CoV-2 neutralizing antibody by wild-type plaque reduction neutralization, microneutralization and pseudotyped virus neutralization assays. *Nat Protoc* 2021; **16**(6): 3114-40.

3.	Mattiuzzo G, Bentley EM, Hassall M, et al. Establishment of the WHO International Standard and Reference Panel for anti-SARS-CoV-2 antibody. *World Health Organization* 2020; **60**.

### 4. Thiruvengadam R, Awasthi A, Medigeshi G Cellular Immune Responses are Preserved and May Contribute to Chadox1 ChAdOx1 nCoV-19 Vaccine Effectiveness Against Infection Due to SARS-CoV-2 B·1·617·2 Delta Variant Despite Reduced Virus Neutralisation. medRxiv <https://doi.org/10.2139/ssrn.3884946>
